# Menopause timing, hormone therapy, epigenetic aging, and breast cancer outcomes: a secondary analysis of the PLCO Cancer Screening Trial

**DOI:** 10.64898/2026.10.04.26364714

**Authors:** Ting Zhai, Vernon S Pankratz, Katherine E Zychowski, Yafang Li, Kimberly K Leslie, Yan Guo, Christopher I Amos, Linda S Cook, Shuguang Leng

## Abstract

**Background:** Current recommendations limit the use of menopausal hormone therapy (MHT) principally to the management of menopausal symptoms, largely because of concerns regarding increased breast cancer (BC) risk. The impact of MHT use on systemic biological aging and blood methylome and its overlap with that of menopause timing remains incompletely understood.

**Methods:** In females (N=78,209) of the Prostate, Lung, Colorectal and Ovarian (PLCO) Cancer Screening Trial, we used Cox models to assess the associations of menopause timing (ordinal age at menopause; binary early-M [<45 years] vs non-early-M [≥45 years]) and MHT use with BC incidence, mortality, and survival. In a nested case-control subset with blood DNA methylome (DNAm; 758 cases, 745 controls), we related both exposures to epigenetic clocks and DNAm-based telomere length (DNAmTL) and used polygenic instruments to test both causal directions.

**Results:** Ever MHT use was associated with higher BC incidence (aHR 1.17, 95% CI 1.10–1.24), particularly for hormone-receptor positive (HR+) subtypes (1.19, 1.11–1.27), whereas early-M was associated with lower HR+ incidence (0.88, 0.81–0.96) but not with overall incidence after adjustment (0.94, 0.88–1.01). However, ever MHT use was associated with lower BC mortality (0.72, 0.62–0.83) and better overall survival (0.87, 0.81–0.95). In the DNAm subset, non-early-M was associated with slower epigenetic aging and longer DNAmTL particularly in cases. Higher cumulative aging and pace-of-aging markers predicted worse survival and partially mediated the early-M associations. Genetic analyses supported shared polygenic architecture between menopause timing and DNAm aging biomarkers rather than simple causation. MHT showed no broad epigenetic age acceleration and its profound methylome and immune-marker signatures largely did not overlap with those of menopause timing.

**Conclusions:** Ever MHT use was associated with lower BC mortality and better overall survival despite higher BC incidence. Systemic aging detected more than a decade after menopausal transition correlates with early menopause. MHT use shows molecular signatures distinct to those seen for menopause timing.

## Introduction

Menopause, the permanent cessation of menstruation, marks the transition to the non-reproductive stage of life and typically occurs between ages 45 and 55 years.^1,2^ After menopause, estrogen is produced primarily by peripheral tissues, particularly adipose tissue.^3^ Menopause may occur naturally or be induced by surgery or medical treatments, and many women use menopausal hormone therapy (MHT) to alleviate symptoms of estrogen withdrawal.^4^ The United States Preventive Services Task Force recommends against MHT for the primary prevention of chronic conditions, whereas the North American Menopause Society (NAMS) supports individualized use in selected women, particularly those younger than 60 years or within 10 years of menopause onset;^4–6^ in November 2025 the United States Food and Drug Administration (FDA) initiated removal of the boxed warnings for cardiovascular disease, breast cancer, and probable dementia from menopausal hormone therapy products, retaining only the endometrial-cancer warning for unopposed estrogen, a shift broadly consistent with the NAMS position.^7^ Because the ovary is among the earliest organs to show functional decline, age at menopause (AAM) reflects not only reproductive status but also the pace of somatic aging, and loci governing ovarian ageing are enriched for DNA damage response and genome maintenance genes.^8–10^ MHT could relate to the same axis from the opposite direction, as estrogen signaling regulates immune and inflammatory function and MHT use has been associated with lower epigenetic age in some tissues.^11,12^

MHT has been reported to slow the progression of biological aging in women.^13^ Epigenetic aging biomarkers derived from DNA methylation (DNAm) profiles quantify biological aging beyond chronological age. First-generation clocks (Horvath, Hannum) primarily capture intrinsic aging, whereas second-generation clocks are more strongly associated with morbidity and mortality because they were trained on aging-related phenotypes rather than chronological age: PhenoAge on a composite of nine clinical chemistry and hematologic markers,^14^ GrimAge on time to death using DNAm surrogates of plasma proteins and smoking pack-years.^15–17^ Telomere length further reflects replicative aging,^18^ and DunedinPACE, trained on the longitudinal rate of decline across 19 organ-system indicators, estimates the current pace rather than the accumulated amount of aging.^19^ Because these metrics are anchored to different training targets, an exposure or outcome need not relate to all of them with the same direction or magnitude, and the pattern across biomarker classes is more informative than any single clock.

Epigenetic age acceleration (EAA), the residual of epigenetic age on chronological age, has been associated with mortality, cancer outcomes, immune dysregulation, and chronic disease risk.^15–17^ Menopause is accompanied by endocrine and immunologic remodeling, raising the possibility that variation in menopause timing is reflected not only in systemic epigenetic aging but also in immune response pathways.^11^ Younger AAM has been linked to lung diseases, cardiovascular diseases, osteoporosis, metabolic disorders, and all-cause mortality,^1,2,8,9,20^ yet studies of menopause timing and epigenetic aging have been inconsistent.^12,21^ Importantly, aging may act as both cause and consequence: accelerated biological aging may predispose women to early-M, whereas early estrogen deprivation may in turn accelerate systemic aging.^12,22^ Disentangling these bidirectional relationships remains challenging in observational studies.

In this study, we leveraged the Prostate, Lung, Colorectal and Ovarian (PLCO) Cancer Screening Trial to evaluate the associations of menopause timing (an ordinal AAM variable and a binary early-M vs non-early-M contrast) and MHT use with BC incidence, mortality, and overall survival, and investigated their relationships with epigenetic aging in a nested BC case-control study.^23^ Causality between age at natural menopause and epigenetic aging was inferred using genetic approaches, and we integrated epigenome-wide association study (EWAS), pathway-enrichment, and immune-marker analyses to test whether the two exposures share biologic signatures. This comparison distinguishes two possibilities: if endogenous and exogenous estrogen exposure act along a common pathway, the two should converge on overlapping methylation, pathway, and immune signatures, whereas divergence would indicate that MHT influences BC outcomes through mechanisms largely separable from systemic aging.

## Methods

### Study Population

Participants were drawn from the Prostate, Lung, Colorectal and Ovarian (PLCO) Cancer Screening Trial (data access: NCI CDAS PLCO-981; dbGaP project 35044), which randomized 154,887 generally healthy, cancer-free adults aged 55–74 years to test whether screening reduces cancer mortality. Clinical analyses used the full female screening cohort (N=78,209), with trial cancer incidence from 1993–2009 and registry-linked follow-up through 2020. Molecular analyses used a nested breast cancer case-control substudy with pre-diagnostic peripheral-blood DNA methylation (Illumina EPIC v1) in 1,568 women (721 incident invasive cases, 847 frequency-matched controls); after methylation-quality exclusions (n=51) and removal of participants missing age at menopause (n=14), the analytic epigenetic cohort comprised 1,503 women, all self-reported White non-Hispanic (Figure 1). AAM was harmonized into five ordinal levels (<40, 40–44, 45–49, 50–54, >=55 years) and dichotomized as early-M (<45 years) versus non-early-M (>=45 years); menopause type and MHT use came from baseline and follow-up questionnaires. Ascertainment, exclusions, and derivations are in the Supplementary Methods. The PLCO trial was approved by the institutional review boards of the National Cancer Institute and the participating screening centers, and all participants provided written informed consent at enrolment.

**Figure 1.**
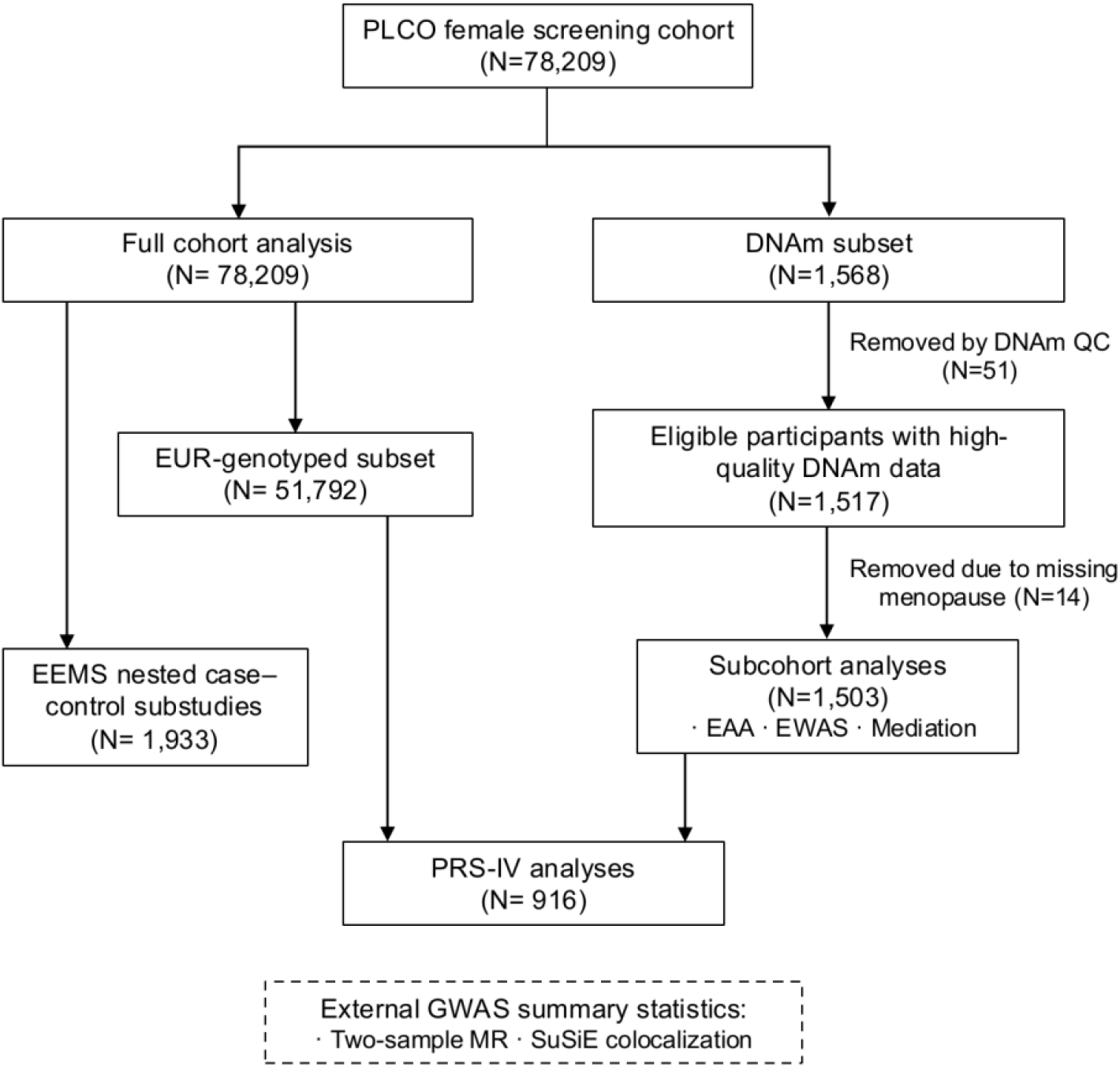
Study design and participant flow. Individual-level analyses used the full PLCO female screening cohort and three nested subsets: participants with genome-wide genotype data, participants in the Etiologic and Early Marker Studies (EEMS) immune substudies, and the breast cancer case–control subset with baseline blood DNA methylation. Polygenic-score instrumental-variable analyses required both methylation and genotype data. The dashed box denotes analyses based solely on published GWAS summary statistics, which included no PLCO participants.

### DNA Methylation and Epigenetic Aging

Raw EPIC signals were processed in minfi^24^ with standard quality control, single-sample Noob (ssNoob) background and dye-bias correction,^25^ and quantile normalization, retaining 824,126 probes; blood cell-type proportions were estimated by reference-based deconvolution. Six DNAm aging metrics were computed via the Clock Foundation platform (https://dnamage.clockfoundation.org): the Horvath, Hannum, PhenoAge, and GrimAge clocks, DNAmTL, and DunedinPACE, with epigenetic age acceleration (EAA) defined as the cell-composition-adjusted residual of each clock on chronological age (Figure S1A). Constituent CpGs, EPIC coverage, and clock derivations are given in the Supplementary Methods.

### Epigenome-wide and pathway analyses

EWAS was performed with limma^26^ empirical-Bayes models on M-values, testing AAM (ordinal) and, in parallel models, MHT use, adjusted for chronological age, BMI, education, MHT/menopause type, cancer status, cell composition, and technical covariates; Benjamini-Hochberg (BH) FDR was applied, with a suggestive threshold of 5×10^−6^ for locus prioritization. Gene-set enrichment used *methylGSA* (mRRA) across MSigDB Hallmark and KEGG pathways.

### Genetic analyses

Among European-ancestry PLCO participants, polygenic risk scores (PRS) for age at natural menopause (ANM) and for EAA were constructed with LDpred2-auto on HapMap3 variants using published GWAS summary statistics external to PLCO (ANM PRS explained 9.9% of ANM variance; Figure S2A), adjusting for the first eight genetic principal components which together explained 67% of total genetic variance (Figure S2D); pairwise PRS correlations used Pearson coefficients. Directional ANM-EAA relationships were evaluated with PRS-based instrumental-variable structural equation models (lavaan) in PLCO individual-level data, a one-sample design, complemented by SNP-level two-sample Mendelian randomization (CAUSE, TwoSampleMR, MR-PRESSO) using non-overlapping exposure and outcome GWAS, and SuSiE-based colocalization of the same GWAS pairs. Source GWAS, genotyping QC, and all specifications are in the Supplementary Methods.

### Immune Marker Analysis

Pre-diagnostic serum immune and inflammation markers were pooled from five nested case-control substudies conducted within PLCO for different cancer sites (EEMS) and restricted to women, yielding 74 QC-retained analytes (Table S1); 48 markers with >=50% non-missing data in the analytic sample (n=1,933) were tested. Associations with AAM (and, in parallel, MHT use) used linear models with source-substudy fixed effects on log-transformed, standardized concentrations, with fixed-effect meta-analysis across substudies to assess heterogeneity (**Table S2**) and BH FDR correction.

### Statistical Analysis

Associations of AAM and MHT use (status, duration, and regimens) with BC incidence, BC-specific mortality, and overall survival among cases were evaluated using age-adjusted (Model 1) and multivariable (Model 2) Cox models. Incidence and mortality analyses used the full female cohort with time from randomization; overall survival was analysed among incident cases only with time from diagnosis. Model 2 adjusted for age, BMI, education, menopause type, smoking, and the alternate exposure. For case-only survival analyses, stage at diagnosis was included in both Model 1 and Model 2. MHT duration was taken from the baseline report of total years of hormone use and reverse-coded as an ordinal exposure (never use, ^≤^1, 2–3, 4–5, 6–9, and ≥10 years); because duration was modelled as an ordinal variable, women missing duration were excluded from duration analyses. MHT regimen was derived from the supplemental questionnaire item on the type of therapy first used, aligned with the baseline questionnaire, and classified as never use, estrogen-only pills, progestin pills, estrogen plus progestin pills, creams or other non-oral preparations, and ever use with unknown regimen; regimen was examined within strata of menopause type. Participants missing a categorical covariate were retained via an explicit unknown category, and competing-/unknown-subtype cases were censored in subtype-stratified analyses. AAM was modeled both as a 5-level ordinal exposure and as the binary early-M contrast. Associations with aging metrics and cell-type proportions used linear models, with effect modification assessed by menopause type, smoking status, and BC status. Mediation was assessed by sequential adjustment and regression-based causal mediation on the multiplicative hazards scale (CMAverse, 2,000 bootstraps), decomposing the total effect into natural direct and total natural indirect effects (TNIE). Tests were two-sided with FDR correction within analytic domains; analyses used R v4.4.1.

## Results

### Study participant characteristics

Among the 78,209 female participants in the PLCO (mean baseline age 62.5 years), 6,721 received a BC diagnosis over 1,262,412 person-years of follow-up; 908 breast cancer deaths occurred cohort-wide over 1,554,945 person-years, and 3,008 of the cases died of any cause over 79,370 person-years of follow-up from diagnosis. AAM was <40 years in 14.5%, 40–44 in 14.1%, 45–49 in 23.8%, 50–54 in 36.4%, and >=55 in 11.1%; 66.1% reported ever MHT use (49.3% current at baseline). The nested DNAm study included 1,503 postmenopausal women (745 controls, 758 BC cases by updated-registry status; Table 1), broadly similar between cases and controls in demographic, lifestyle, and reproductive characteristics, so epigenetic analyses were conducted in the combined sample with BC status accounted for analytically. This is consistent with a meta-analysis of four prospective cohorts (1,663 cases, 1,885 controls) reporting no association between blood methylation at individual CpGs and subsequent BC risk.^23^

**Table 1.** Cohort characteristics by breast cancer status in EPIC methylation subset and full PLCO female cohort.

| Characteristic | EPIC subset |  |  |  |  | Full PLCO cohort |  |  |  |  |
| --- | --- | --- | --- | --- | --- | --- | --- | --- | --- | --- |
|  | All (n=1,503) | Control (n=745) | Breast cancer (n=758) | P value | % missing | Overall (n=78,209) | No breast cancer (n=71,488) | Breast cancer (n=6,721) | P value | % missing |
| n | 1503 | 745 | 758 |  |  | 78209 | 71488 | 6721 |  |  |
| Age, years (mean (SD)) | 62.01 (5.21) | 62.09 (5.16) | 61.92 (5.26) | 0.533 | 0.0 | 62.51 (5.40) | 62.54 (5.41) | 62.25 (5.25) | <0.001 | 0.0 |
| Pack-years (mean (SD)) | 10.60 (19.09) | 9.89 (17.83) | 11.30 (20.25) | 0.152 | 0.5 | 13.25 (22.46) | 13.27 (22.47) | 13.14 (22.30) | 0.661 | 3.6 |
| Race/ethnicity, n (%) |  |  |  |  | 0.0 |  |  |  | <0.001 | 0.0 |
| White, Non-Hispanic | 1503 (100) | 745 (100) | 758 (100) |  |  | 67395 (86.2) | 61459 (86.0) | 5936 (88.3) |  |  |
| Black, Non-Hispanic | 0 (0) | 0 (0) | 0 (0) |  |  | 4338 (5.5) | 4092 (5.7) | 246 (3.7) |  |  |
| Hispanic | 0 (0) | 0 (0) | 0 (0) |  |  | 1215 (1.6) | 1133 (1.6) | 82 (1.2) |  |  |
| Asian | 0 (0) | 0 (0) | 0 (0) |  |  | 2567 (3.3) | 2324 (3.3) | 243 (3.6) |  |  |
| Pacific Islander | 0 (0) | 0 (0) | 0 (0) |  |  | 371 (0.5) | 326 (0.5) | 45 (0.7) |  |  |
| American Indian | 0 (0) | 0 (0) | 0 (0) |  |  | 202 (0.3) | 194 (0.3) | 8 (0.1) |  |  |
| Missing | 0 (0) | 0 (0) | 0 (0) |  |  | 2121 (2.7) | 1960 (2.7) | 161 (2.4) |  |  |
| Hispanic ethnicity, n (%) | 0 (0) | 0 (0) | 0 (0) |  | 0.0 | 1302 (1.8) | 1216 (1.8) | 86 (1.3) | 0.012 | 5.0 |
| Education, n (%) |  |  |  | 0.224 | 0.0 |  |  |  | <0.001 | 2.9 |
| Less than high school | 69 (4.6) | 37 (5.0) | 32 (4.2) |  |  | 4985 (6.6) | 4671 (6.7) | 314 (4.8) |  |  |
| High school graduate | 410 (27.3) | 216 (29.0) | 194 (25.6) |  |  | 20910 (27.5) | 19255 (27.8) | 1655 (25.3) |  |  |
| Some college or higher | 1024 (68.1) | 492 (66.0) | 532 (70.2) |  |  | 50022 (65.9) | 45446 (65.5) | 4576 (69.9) |  |  |
| Married, n (%) | 1141 (76.0) | 568 (76.3) | 573 (75.6) | 0.780 | 0.1 | 52275 (68.8) | 47666 (68.7) | 4609 (70.4) | 0.005 | 2.9 |
| Smoking status, n (%) |  |  |  | 0.266 | 0.0 |  |  |  | <0.001 | 2.7 |
| Never | 905 (60.2) | 464 (62.3) | 441 (58.2) |  |  | 42352 (55.6) | 38772 (55.8) | 3580 (54.6) |  |  |
| Former | 497 (33.1) | 234 (31.4) | 263 (34.7) |  |  | 26359 (34.6) | 23908 (34.4) | 2451 (37.4) |  |  |
| Current | 101 (6.7) | 47 (6.3) | 54 (7.1) |  |  | 7395 (9.7) | 6865 (9.9) | 530 (8.1) |  |  |
| BMI category, n (%) |  |  |  | 0.948 | 0.9 |  |  |  | 0.008 | 4.1 |
| Underweight (<18.5 kg/m<sup>2</sup>) | 13 (0.9) | 6 (0.8) | 7 (0.9) |  |  | 863 (1.2) | 814 (1.2) | 49 (0.8) |  |  |
| Normal weight (18.5 to <25 kg/m<sup>2</sup>) | 570 (38.3) | 285 (38.6) | 285 (37.9) |  |  | 30084 (40.1) | 27533 (40.2) | 2551 (39.4) |  |  |
| Overweight (25 to <30 kg/m<sup>2</sup>) | 548 (36.8) | 267 (36.1) | 281 (37.4) |  |  | 25773 (34.4) | 23500 (34.3) | 2273 (35.1) |  |  |
| Obese (≥30 kg/m<sup>2</sup>) | 359 (24.1) | 181 (24.5) | 178 (23.7) |  |  | 18297 (24.4) | 16695 (24.4) | 1602 (24.7) |  |  |
| Menopause type, n (%) |  |  |  | 0.85 | 1.3 |  |  |  | <0.001 | 4.5 |
| Natural | 917<br>(61.8) | 458<br>(62.1) | 459<br>(61.5) |  |  | 46,337<br>(62.0) | 42,274<br>(61.9) | 4,063<br>(63.2) |  |  |
| Surgical | 501<br>(33.8) | 249<br>(33.8) | 252<br>(33.8) |  |  | 25,638<br>(34.3) | 23,610<br>(34.6) | 2,028<br>(31.6) |  |  |
| Radiation | 0 (0) | 0 (0) | 0 (0) |  |  | 107 (0.1) | 99 (0.1) | 8 (0.1) |  |  |
| Drug-induced | 65 (4.4) | 30 (4.1) | 35 (4.7) |  |  | 2,626 (3.5) | 2,301 (3.4) | 325 (5.1) |  |  |
| Age at<br>menopause /<br>last<br>menstrual<br>period, n (%) |  |  |  | 0.66<br>2 | 0.0 |  |  |  | <0.00<br>1 | 3.5 |
| <40 years | 177<br>(11.8) | 79 (10.6) | 98 (12.9) |  |  | 10922<br>(14.5) | 10100<br>(14.6) | 822 (12.7) |  |  |
| 40–44 years | 228<br>(15.2) | 111<br>(14.9) | 117<br>(15.4) |  |  | 10652<br>(14.1) | 9804 (14.2) | 848 (13.1) |  |  |
| 45–49 years | 331<br>(22.0) | 164<br>(22.0) | 167<br>(22.0) |  |  | 17993<br>(23.8) | 16530<br>(24.0) | 1463<br>(22.5) |  |  |
| 50–54 years | 555<br>(36.9) | 283<br>(38.0) | 272<br>(35.9) |  |  | 27507<br>(36.4) | 24998<br>(36.2) | 2509<br>(38.6) |  |  |
| ≥55 years | 212<br>(14.1) | 108<br>(14.5) | 104<br>(13.7) |  |  | 8397 (11.1) | 7545 (10.9) | 852 (13.1) |  |  |
| Age at first<br>menstrual<br>period, n (%) |  |  |  | 0.85<br>3 | 0.1 |  |  |  | 0.243 | 2.9 |
| <10 years | 13 (0.9) | 7 (0.9) | 6 (0.8) |  |  | 1236 (1.6) | 1132 (1.6) | 104 (1.6) |  |  |
| 10–11 years | 284<br>(18.9) | 141<br>(19.0) | 143<br>(18.9) |  |  | 14219<br>(18.7) | 12925<br>(18.6) | 1294<br>(19.8) |  |  |
| 12–13 years | 818<br>(54.5) | 411<br>(55.2) | 407<br>(53.7) |  |  | 40797<br>(53.7) | 37314<br>(53.8) | 3483<br>(53.2) |  |  |
| 14–15 years | 330<br>(22.0) | 155<br>(20.8) | 175<br>(23.1) |  |  | 16201<br>(21.3) | 14824<br>(21.4) | 1377<br>(21.0) |  |  |
| ≥16 years | 57 (3.8) | 30 (4.0) | 27 (3.6) |  |  | 3451 (4.5) | 3166 (4.6) | 285 (4.4) |  |  |
| Ever use oral<br>contraceptive<br>s, n (%) | 869<br>(57.8) | 438<br>(58.8) | 431<br>(56.9) | 0.48<br>0 | 0.0 | 41195<br>(54.2) | 37591<br>(54.1) | 3604<br>(55.0) | 0.167 | 2.9 |
| Hormone<br>therapy use,<br>n (%) |  |  |  | 0.15<br>0 | 0.2 |  |  |  | <0.00<br>1 | 3.4 |
| Never | 427<br>(28.5) | 215<br>(29.0) | 212<br>(28.0) |  |  | 25439<br>(33.7) | 23509<br>(34.0) | 1930<br>(29.6) |  |  |
| Former | 211<br>(14.1) | 116<br>(15.6) | 95 (12.5) |  |  | 12854<br>(17.0) | 11830<br>(17.1) | 1024<br>(15.7) |  |  |
| Current | 862<br>(57.5) | 411<br>(55.4) | 451<br>(59.5) |  |  | 37288<br>(49.3) | 33714<br>(48.8) | 3574<br>(54.7) |  |  |
| Ever<br>hormone<br>therapy use,<br>n (%) | 1073<br>(71.4) | 527<br>(70.7) | 546<br>(72.0) | 0.61<br>9 | 0.0 | 50243<br>(66.1) | 45641<br>(65.7) | 4602<br>(70.3) | <0.00<br>1 | 2.8 |
| Hysterectomy<br>, n (%) | 532<br>(35.4) | 262<br>(35.2) | 270<br>(35.6) | 0.89<br>7 | 0.0 | 27575<br>(36.3) | 25369<br>(36.5) | 2206<br>(33.7) | <0.00<br>1 | 2.8 |
| Bilateral<br>oophorectom<br>y, n (%) | 284<br>(18.9) | 143<br>(19.2) | 141<br>(18.6) | 0.82<br>0 | 0.0 | 15342<br>(19.6) | 14140<br>(19.8) | 1202<br>(17.9) | <0.00<br>1 | 0.0 |
| Breast cancer<br>family history,<br>n (%) | 239<br>(16.0) | 98 (13.2) | 141<br>(18.8) | 0.00<br>4 | 0.9 | 10696<br>(14.1) | 9415 (13.6) | 1281<br>(19.7) | <0.00<br>1 | 3.3 |
| Number of<br>pregnancies<br>(mean (SD)) | 2.77<br>(1.15) | 2.80<br>(1.16) | 2.75<br>(1.15) | 0.36<br>8 | 0.1 | 2.78 (1.18) | 2.79 (1.17) | 2.70 (1.20) | <0.00<br>1 | 2.9 |

Abbreviations: BMI=body mass index; SD=standard deviation.

### MHT use is associated with higher incidence but lower breast cancer mortality

In the full PLCO female cohort, early-M was inversely associated with BC incidence in age-adjusted models (HR 0.89, 95% CI 0.84–0.94), but this association was attenuated after covariate adjustment (aHR 0.94, 95% CI 0.88–1.01, Table 2), reflecting collinearity with surgical menopause, which both lowers BC risk and accounts for the majority of early-M cases; when menopause type was removed from the model, the early-M association strengthened to aHR 0.88 (95% CI 0.83–0.93) in a complete-case analysis. For BC-specific mortality, early-M showed a non-significant inverse association (aHR 0.86, 95% CI 0.71–1.04). Ordinal AAM results were directionally consistent (Table S3).

**Table 2.** Early menopause, ever hormone therapy use, and duration of hormone therapy use in relation to breast cancer incidence, breast cancer-specific mortality, and overall survival among cases.

| Exposure | Endpoint | N (Mod 1) | Events (Mod 1) | Model 1 HR (95% CI) | Model 1 P | N (Mod 2) | Events (Mod 2) | Model 2 HR (95% CI) | Model 2 P |
| --- | --- | --- | --- | --- | --- | --- | --- | --- | --- |
| Early menopause (<45 vs ≥45 years) | BC incidence | 75471 | 6494 | **0.89 (0.84–0.94)** | **<0.001** | 75471 | 6494 | 0.94 (0.88–1.01) | 0.077 |
| Early menopause (<45 vs ≥45 years) | BC mortality | 75471 | 872 | 0.88 (0.75–1.02) | 0.090 | 75471 | 872 | 0.86 (0.71–1.04) | 0.116 |
| Early menopause (<45 vs ≥45 years) | OS (cases) | 6494 | 2901 | 1.09 (1.00–1.18) | 0.050 | 6494 | 2901 | 1.07 (0.96–1.18) | 0.219 |
| Ever MHT use (vs never) | BC incidence | 75682 | 6532 | **1.16 (1.10–1.23)** | **<0.001** | 75682 | 6532 | **1.17 (1.10–1.24)** | **<0.001** |
| Ever MHT use (vs never) | BC mortality | 75682 | 869 | **0.73 (0.63–0.84)** | **<0.001** | 75682 | 869 | **0.72 (0.62–0.83)** | **<0.001** |
| Ever MHT use (vs never) | OS (cases) | 6532 | 2911 | **0.82 (0.76–0.89)** | **<0.001** | 6532 | 2911 | **0.87 (0.81–0.95)** | **0.001** |
| MHT duration (per category increase) | BC incidence | 75467 | 6518 | **1.04 (1.03–1.05)** | **<0.001** | 75467 | 6518 | **1.05 (1.03–1.06)** | **<0.001** |
| MHT duration (per category increase) | BC mortality | 75467 | 869 | **0.95 (0.92–0.98)** | **0.003** | 75467 | 869 | **0.95 (0.91–0.98)** | **0.004** |
| MHT duration (per category increase) | OS (cases) | 6518 | 2903 | **0.96 (0.94–0.98)** | **<0.001** | 6518 | 2903 | **0.97 (0.95–0.99)** | **0.002** |

Ever MHT use was associated with higher BC incidence (aHR 1.17, 95% CI 1.10–1.24), lower BC-specific mortality (aHR 0.72, 95% CI 0.62–0.83), and improved overall survival among cases (aHR 0.87, 95% CI 0.81–0.95; Table 2). MHT regimens differed substantially by menopause type. Among women with natural menopause, combined estrogen-progestin therapy (47.5%) was more common than estrogen-only therapy (34.9%), whereas estrogen-only therapy predominated among women with surgical menopause (85.2%), consistent with the very high prevalence of hysterectomy (99.7%) in this group. By subtype, the MHT-associated increase was restricted to HR+ tumors (aHR 1.19, 95% CI 1.11–1.27; HR-1.03, 0.87–1.21), as was the early-M reduction (HR+ 0.88, 0.81–0.96; HR-1.03, 0.84–1.26); subtype heterogeneity was significant for early-M (p=0.032) but not MHT use (p=0.23) (Table S5). Longer MHT use was associated with progressively higher BC incidence (aHR 1.05, 1.03–1.06 per one-category increase in duration, p<0.001), and with lower BC-specific mortality (0.95, 0.91–0.98, p=0.004) and better overall survival (0.97, 0.95–0.99, p=0.002; Table 2). Associations differed by menopause type (Table S4): the incidence excess with ever MHT use was confined to women with natural menopause (1.22, 1.15–1.31) and was absent after surgical menopause (1.05, 0.94–1.18; p for interaction 0.018). Among women with natural menopause, the excess was larger for estrogen plus progestin pills (1.29, 1.17–1.42, p<0.001) than for estrogen-only pills (1.13, 1.02–1.26, p=0.021), with ever users of unknown regimen intermediate (1.23, 1.14–1.33). The lower BC-specific mortality was seen with both regimens (estrogen-only 0.51, 0.36–0.72; estrogen plus progestin 0.60, 0.45–0.81).

### Earlier menopause is associated with epigenetic age acceleration

Menopause occurred a mean of 13.5 years before blood collection, so these associations reflect DNAm differences persisting well beyond the transition (Table S6). Because age at collection was similar across AAM groups (means 61.5–63.6 years), time since menopause is essentially the inverse of AAM and is accounted for by adjustment for age at collection. Earlier menopause was associated with higher HannumAgeAccel and GrimAgeAccel, strongest for GrimAgeAccel (adjusted beta = 0.355 per step earlier AAM, 95% CI 0.154–0.557); associations with PhenoAgeAccel, faster DunedinPACE, and shorter DNAmTL were directionally consistent but did not reach significance (p=0.054–0.074). All five were more pronounced among BC cases, where DunedinPACE and DNAmTL reached significance (Table S7); results were consistent for early-M versus non-early-M. This pattern was driven by natural menopause (GrimAgeAccel beta = 0.480, 95% CI 0.192–0.767) and attenuated in non-natural menopause (0.212, -0.032 to 0.456), whereas DNAmTL was not associated in either stratum. In contrast, MHT use showed little association with epigenetic aging; AAM was unrelated to DNAm-estimated cell composition, whereas current MHT use was linked to lower B-cell and higher granulocyte proportions (Table S8; Figure S3).

### Epigenetic aging partly mediates early menopause and survival

None of the six aging biomarkers differed between BC cases and controls (Table S9). Among 758 BC cases with DNAm data, higher DunedinPACE (aHR 1.47, 95% CI 1.28–1.69) and GrimAgeAccel (aHR 1.41, 95% CI 1.22–1.63) were the strongest predictors of poorer overall survival (Table 3). Early-M was associated with worse survival (aHR 1.38, 95% CI 1.01–1.89), attenuated after adjustment for GrimAgeAccel or DunedinPACE, and mediation analyses identified significant indirect effects through both GrimAgeAccel (total natural indirect effect [TNIE] HR 1.08, 95% CI 1.02–1.19) and DunedinPACE (TNIE HR 1.09, 95% CI 1.02–1.19), supporting partial mediation by accelerated aging.

**Table 3.**
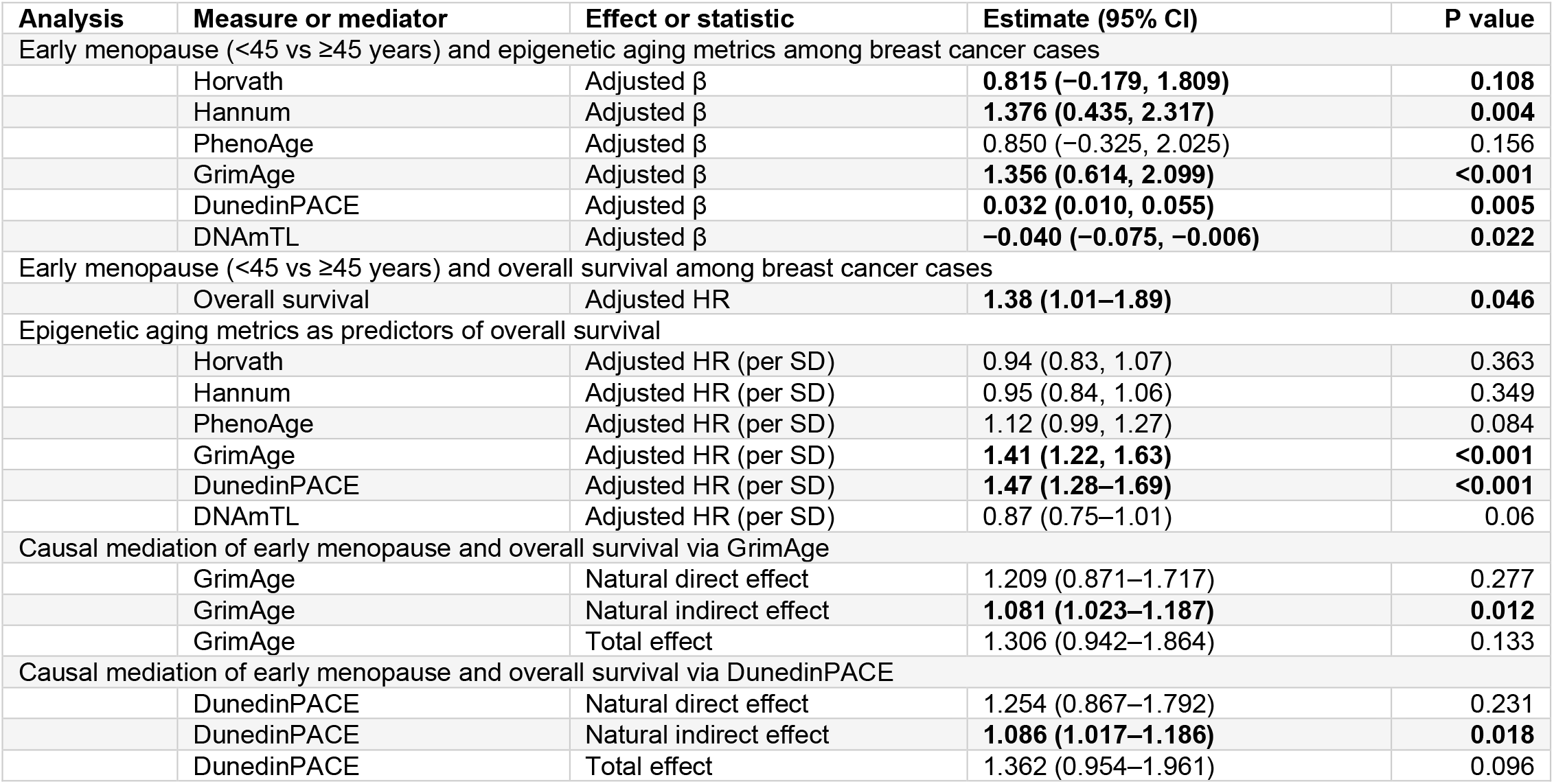
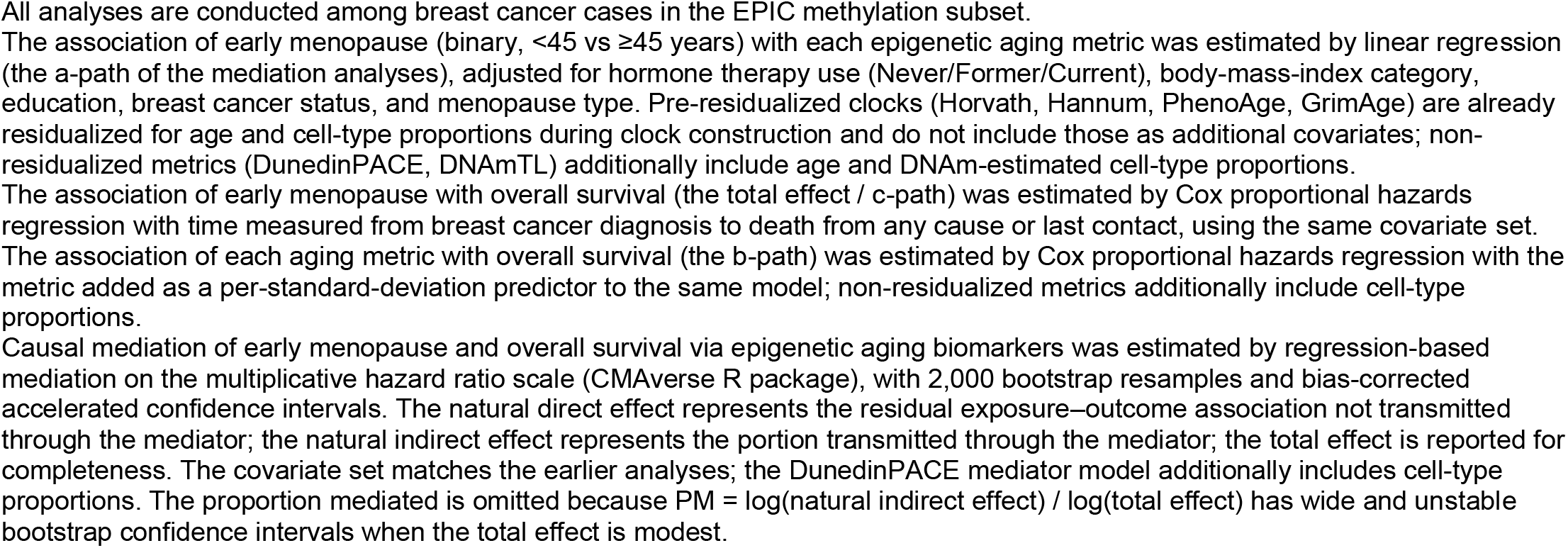
Mediation of early menopause and overall survival via epigenetic aging.

| Analysis | Measure or mediator | Effect or statistic | Estimate (95% CI) | P value |
| --- | --- | --- | --- | --- |
| Early menopause (<45 vs ≥45 years) and epigenetic aging metrics among breast cancer cases |  |  |  |  |
|  | Horvath | Adjusted β | **0.815 (–0.179, 1.809)** | **0.108** |
|  | Hannum | Adjusted β | **1.376 (0.435, 2.317)** | **0.004** |
|  | PhenoAge | Adjusted β | 0.850 (–0.325, 2.025) | 0.156 |
|  | GrimAge | Adjusted β | **1.356 (0.614, 2.099)** | **<0.001** |
|  | DunedinPACE | Adjusted β | **0.032 (0.010, 0.055)** | **0.005** |
|  | DNAmTL | Adjusted β | **–0.040 (–0.075, –0.006)** | **0.022** |
| Early menopause (<45 vs ≥45 years) and overall survival among breast cancer cases |  |  |  |  |
|  | Overall survival | Adjusted HR | **1.38 (1.01–1.89)** | **0.046** |
| Epigenetic aging metrics as predictors of overall survival |  |  |  |  |
|  | Horvath | Adjusted HR (per SD) | 0.94 (0.83, 1.07) | 0.363 |
|  | Hannum | Adjusted HR (per SD) | 0.95 (0.84, 1.06) | 0.349 |
|  | PhenoAge | Adjusted HR (per SD) | 1.12 (0.99, 1.27) | 0.084 |
|  | GrimAge | Adjusted HR (per SD) | **1.41 (1.22, 1.63)** | **<0.001** |
|  | DunedinPACE | Adjusted HR (per SD) | **1.47 (1.28–1.69)** | **<0.001** |
|  | DNAmTL | Adjusted HR (per SD) | 0.87 (0.75–1.01) | 0.06 |
| Causal mediation of early menopause and overall survival via GrimAge |  |  |  |  |
|  | GrimAge | Natural direct effect | 1.209 (0.871–1.717) | 0.277 |
|  | GrimAge | Natural indirect effect | **1.081 (1.023–1.187)** | **0.012** |
|  | GrimAge | Total effect | 1.306 (0.942–1.864) | 0.133 |
| Causal mediation of early menopause and overall survival via DunedinPACE |  |  |  |  |
|  | DunedinPACE | Natural direct effect | 1.254 (0.867–1.792) | 0.231 |
|  | DunedinPACE | Natural indirect effect | **1.086 (1.017–1.186)** | **0.018** |
|  | DunedinPACE | Total effect | 1.362 (0.954–1.961) | 0.096 |

All analyses are conducted among breast cancer cases in the EPIC methylation subset.
The association of early menopause (binary, <45 vs ≥45 years) with each epigenetic aging metric was estimated by linear regression (the a-path of the mediation analyses), adjusted for hormone therapy use (Never/Former/Current), body-mass-index category, education, breast cancer status, and menopause type. Pre-residualized clocks (Horvath, Hannum, PhenoAge, GrimAge) are already residualized for age and cell-type proportions during clock construction and do not include those as additional covariates; non-residualized metrics (DunedinPACE, DNAmTL) additionally include age and DNAm-estimated cell-type proportions.
The association of early menopause with overall survival (the total effect / c-path) was estimated by Cox proportional hazards regression with time measured from breast cancer diagnosis to death from any cause or last contact, using the same covariate set.
The association of each aging metric with overall survival (the b-path) was estimated by Cox proportional hazards regression with the metric added as a per-standard-deviation predictor to the same model; non-residualized metrics additionally include cell-type proportions.
Causal mediation of early menopause and overall survival via epigenetic aging biomarkers was estimated by regression-based mediation on the multiplicative hazard ratio scale (CMAverse R package), with 2,000 bootstrap resamples and bias-corrected accelerated confidence intervals. The natural direct effect represents the residual exposure–outcome association not transmitted through the mediator; the natural indirect effect represents the portion transmitted through the mediator; the total effect is reported for completeness. The covariate set matches the earlier analyses; the DunedinPACE mediator model additionally includes cell-type proportions. The proportion mediated is omitted because $PM = \log(\text{natural indirect effect}) / \log(\text{total effect})$ has wide and unstable bootstrap confidence intervals when the total effect is modest.

### Menopause timing and epigenetic aging share polygenic architecture

To investigate the genetic relationship between age at natural menopause (ANM) and epigenetic aging, we constructed polygenic risk scores (PRSs) using large independent GWAS summary statistics, prioritizing genome-wide PRS analyses because individual menopause-associated variants have small and often pleiotropic effects. Analyses focused on ANM, as genetic instruments are specific to natural menopause; PRSs were available only for HannumAgeAccel, PhenoAgeAccel, and GrimAgeAccel.

At the polygenic level, the ANM PRS was inversely correlated with PRSs for all three epigenetic age acceleration (EAA) measures (Figure 2A), indicating that individuals genetically predisposed to later menopause also tended to have lower genetically predicted biological aging, concordant with the observational findings and motivating instrumental variable (IV) analyses.

**Figure 2.**
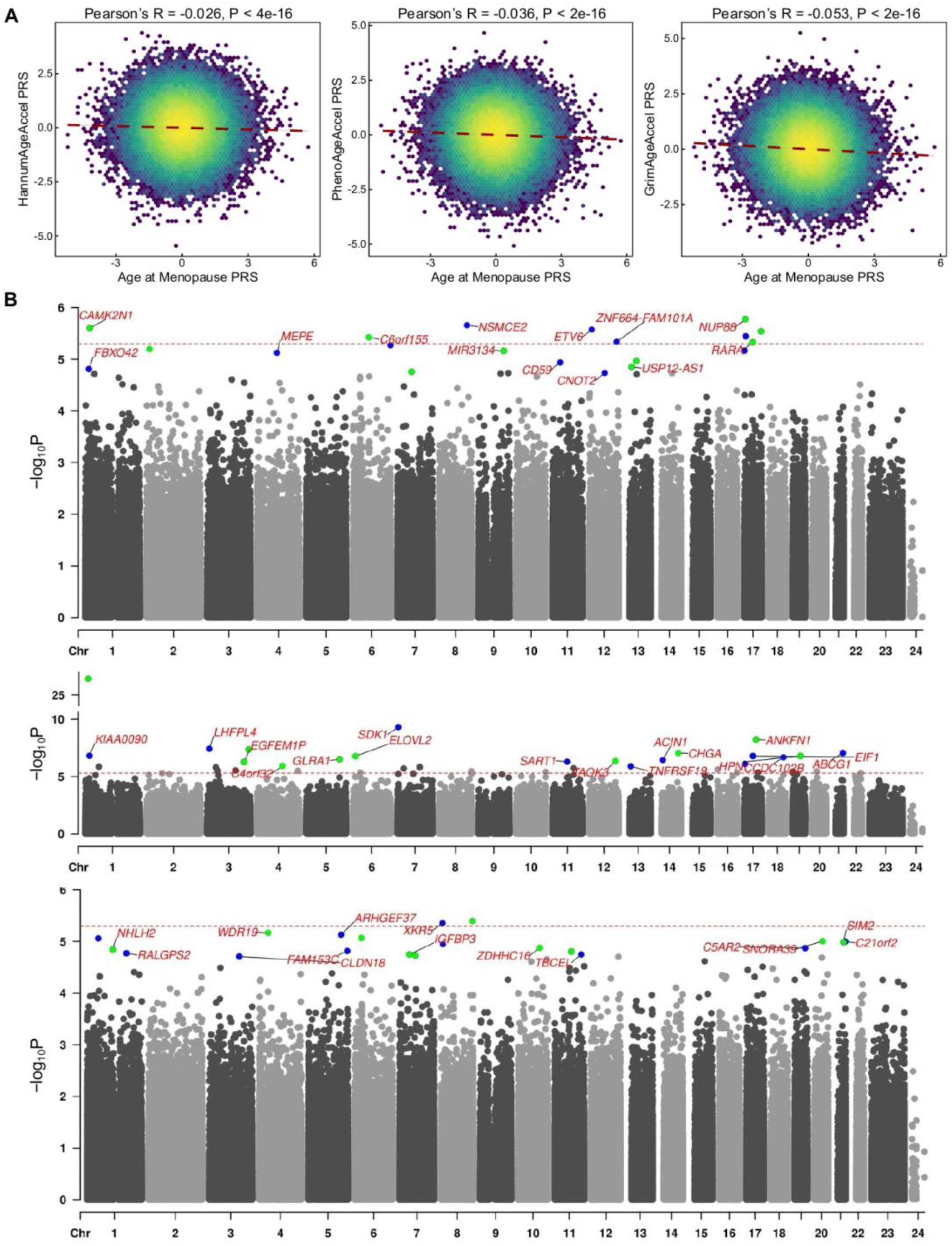
Genetic instruments and epigenome-wide association analyses for menopause timing and HT use. **(A)** Correlation structure between polygenic risk scores (PRS) for age at natural menopause (ANM) and PRSs for epigenetic age acceleration measures (HannumAgeAccel, PhenoAgeAccel, GrimAgeAccel). Correlations are Pearson coefficients computed using standardized PRSs (mean 0, SD 1); higher ANM PRS corresponds to genetically predicted later menopause. **(B)** Manhattan plots from separate EWAS of age at menopause (top) and current (middle) and former (bottom) HT use using limma with empirical Bayes shrinkage. Models adjust for chronological age, BMI, education, HT use/menopause timing, menopause type, cancer diagnosis, blood cell-type proportions, and technical covariates (slide, position). The horizontal line indicates the prespecified suggestive threshold (p<5×10^-6^) and multiple testing is controlled using BH FDR across probes.

In forward PRS-based IV analyses (Table 4), genetically predicted later ANM was associated with lower HannumAgeAccel, PhenoAgeAccel, GrimAgeAccel, and DunedinPACE, as well as longer DNAm-estimated telomere length (DNAmTL), directionally consistent with phenotype-based analyses, whereas associations with HorvathAgeAccel were weaker and non-significant. Estimates represent one-standard-deviation increases in the PRS-explained portion of ANM rather than in the raw PRS, and are therefore not directly comparable in magnitude with ordinal phenotype-based estimates. Pleiotropy-robust sensitivity analyses attenuated HannumAgeAccel and PhenoAgeAccel but left GrimAgeAccel and DNAmTL robust (Table S10).

**Table 4.** PRS-based instrumental-variable analyses of age at natural menopause and epigenetic aging metrics.

| Epigenetic aging metric | ANM to metric (IV: ANM PRS) | Metric to ANM (IV: metric PRS) |
| --- | --- | --- |
| Horvath age acceleration | $-0.04 \pm 0.032$, $p=0.222$ | PRS unavailable |
| Hannum age acceleration | $-0.062 \pm 0.032$, $p=0.048$ | $-0.076 \pm 0.033$, $p=0.021$ |
| PhenoAge acceleration | $-0.088 \pm 0.033$, $p=0.007$ | $-0.089 \pm 0.038$, $p=0.018$ |
| GrimAge acceleration | $-0.103 \pm 0.033$, $p=0.002$ | $-0.122 \pm 0.037$, $p=0.001$ |
| DunedinPACE | $-0.066 \pm 0.031$, $p=0.030$ | PRS unavailable |
| DNAmTL | $0.087 \pm 0.031$, $p=0.005$ | PRS unavailable |

Estimates are PRS-based instrumental-variable effects ( $\beta \pm SE$ ) from SEM, per 1-SD increase in genetic liability, with the exclusion restriction enforced (direct PRS→outcome path fixed to 0); restricted to European-ancestry women with natural menopause and adjusted for PC1–PC8. "PRS unavailable" = no validated EAA PRS for that metric. ANM, age at natural menopause; DNAmTL, DNAm-estimated telomere length; EAA, epigenetic age acceleration; IV, instrumental variable; PC, principal component; PRS, polygenic risk score; SD, standard deviation; SE, standard error; SEM, structural equation modeling.

Reverse-direction IV analyses showed that higher genetically predicted HannumAgeAccel, PhenoAgeAccel, and GrimAgeAccel were associated with earlier menopause, suggesting shared genetic determinants.

Colocalization generally favored distinct rather than shared causal variants, although strong colocalization was observed for ANM and HannumAgeAccel at a chromosome 6 locus (PP.H4 = 0.897; Table S11); given the smaller EAA GWAS, colocalization was underpowered, so the H3-predominant pattern does not exclude shared causal variants. Two-sample Mendelian randomization likewise provided limited evidence for a directional causal effect of ANM on EAA or vice versa (Table S12).

Overall, the genetic analyses support substantial genome-wide overlap between menopause timing and epigenetic aging without support for a directional causal pathway: significance of polygenic instruments in both directions is itself a signature of sharing, whereas pleiotropy-robust SNP-level analyses were null, a pattern characteristic of correlated pleiotropy.

### Current MHT use has a distinct blood methylation signature

In the EWAS of AAM (Figure 2B, top panel), no CpGs reached FDR significance, although several loci exceeded the suggestive threshold (p<5×10^−6^; Table S13), distributed across chromosomes with top-ranked CpGs in CAMK2N1, NSMCE2, ETV6, ZNF664, and NUP88. Pathway analysis showed modest enrichment of KEGG base excision repair, peroxisome, and amino acid metabolism terms, and Hallmark xenobiotic metabolism, complement, TGF-beta signaling, and apical junction terms (Figure 3A, left panel).

**Figure 3.**
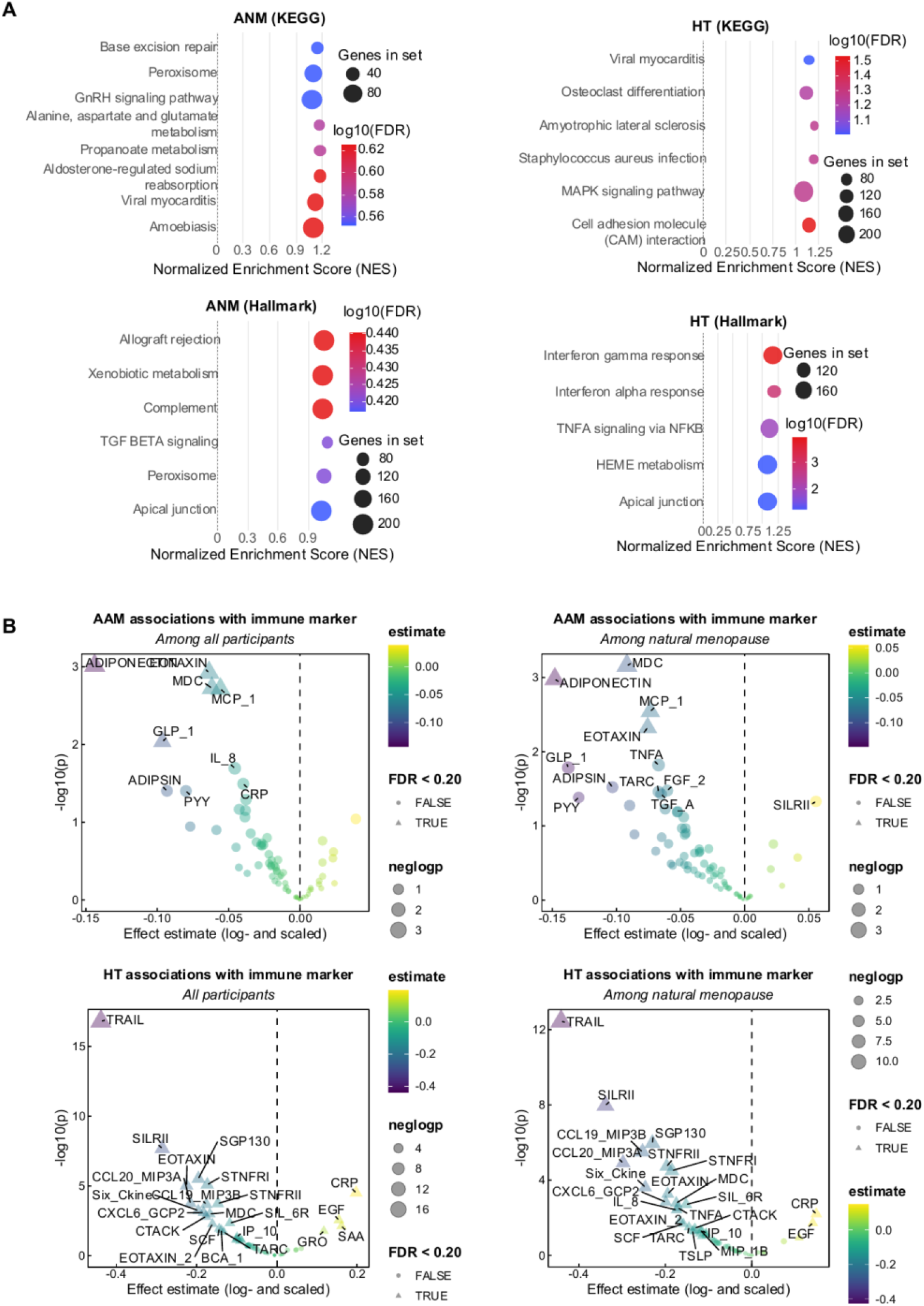
Gene set enrichment and immune marker associations with age at menopause and hormone therapy use. **(A)** Gene set enrichment results based on EWAS-derived CpG signals for menopause timing (left) and HT use (right), including MSigDB Hallmark and KEGG pathways. Enrichment was conducted using methylGSA (mRRA fgsea framework) to account for CpG–gene mapping and probe-count bias; pathway significance is shown as FDR-adjusted results (displayed as NES/FDR per the figure). **(B)** Associations between age at menopause (AAM) and circulating immune or inflammation markers pooled from five PLCO EEMS case-control studies (female participants). Marker concentrations were log-transformed and standardized. Regression models adjust for age, BMI, and study type, with additional adjustment for HT use (AAM models) or AAM (HT models). Results are presented for all participants and for the natural menopause subgroup, with effect estimates shown per ordinal step increase in AAM or contrast between current HT and never users.

EWAS of current versus never MHT use (Figure 2B, middle panel) identified widespread methylation differences (18 CpGs at FDR < 0.05 and 37 at p<5×10^−6^; Table S14), with prominent signals mapping to SDK1, ACIN1, CHGA, ABCG1, EIF1, and ANKFN1, and stronger enrichment than for AAM, highlighting KEGG immune and inflammatory terms (viral myocarditis, MAPK signaling, cell adhesion molecules) and Hallmark interferon-alpha/gamma, TNFalpha/NF-kB, heme metabolism, and epithelial integrity terms (Figure 3A, right panel).

In contrast, the former-versus-never MHT analysis identified no FDR-significant CpGs (two CpGs at p<5×10^−6^; Figure 2B, bottom panel). Nevertheless, five of the 18 FDR-significant CpGs from the current-MHT analysis were nominally associated in former users with concordant but attenuated effect sizes (slope = 0.28, p=0.0008; Figure S1C), a pattern extending genome-wide (slope = 0.59, p<0.001) and indicating a persistent but attenuated methylation signature.

Genome-wide comparison of AAM and current-MHT EWAS results showed virtually no correlation in effect sizes (Pearson r = -0.01), and although 26 of 46 suggestive CpGs (57%) showed concordant directions, the findings support distinct rather than convergent epigenomic signatures.

### MHT and menopause timing have distinct immune-marker profiles

We next examined circulating immune and inflammatory markers (Figure 3B; Table S2). Associations with AAM were modest, earlier menopause tending toward higher EOTAXIN, MCP-1, IL-8, and CRP. Current MHT use showed broader differences: lower TRAIL, EOTAXIN, CCL21 (Six_Ckine), CCL19, and CCL20, and higher CRP and SAA. TRAIL showed substantial between-substudy heterogeneity (I2 = 63%, p=0.045), whereas CRP, SAA, CCL21, CCL19, and CCL20 were highly consistent across the five contributing substudies (I2 = 0%). Across the 48 markers, AAM and current-MHT estimates were weakly and non-significantly correlated (r = -0.13, 95% CI -0.40 to 0.16; p=0.39; r = 0.03 among the 25 markers measured in all five substudies).

## Discussion

We examined associations of AAM and ever MHT use with BC incidence, mortality, and survival in the full PLCO cohort, and whether menopause timing is reflected in DNAm-based aging measures in a nested case-control subset. Later AAM and ever MHT use were each associated with increased BC incidence, particularly HR+ subtypes, consistent with the lifetime cumulative-estrogen-exposure hypothesis for HR+ tumors and the relative estrogen-independence of HR-biology. Neither was associated with an increased likelihood of BC death; both were associated with prolonged survival, with ever MHT use associated with almost 30% lower BC-specific mortality. The increase in incidence was confined to women with natural menopause and was greater for estrogen plus progestin than for estrogen-only therapy, whereas the lower BC-specific mortality was seen with both regimens. HR+ BCs have a more favorable prognosis and greater endocrine-therapy responsiveness, and previous studies, including ours, have linked greater lifetime estrogen exposure with lower all-cause mortality.^1,2,27^ The favorable prognosis may therefore reflect both tumor composition and broader health benefits of prolonged estrogen exposure.

The DNAm analysis extends this clinical pattern: earlier menopause was associated with GrimAgeAccel and HannumAgeAccel, with directionally consistent but non-significant associations for the other metrics, consistent with earlier work linking menopause timing to epigenetic age acceleration.^12,15–17^ Although these measures did not differ between cases and controls, their associations with earlier menopause were primarily driven by those who subsequently developed BC, suggesting that epigenetic aging is more sensitive to estrogen exposure in women in whom BC later develops. DunedinPACE and GrimAgeAccel were the strongest predictors of poorer survival and attenuated the early-M association, compatible with partial mediation. The pattern across biomarkers matches what each metric was designed to measure: GrimAgeAccel and DunedinPACE, which encode inflammatory and exposure-related burden and the current rate of multisystem decline, predicted survival strongly, whereas HorvathAgeAccel and HannumAgeAccel, calibrated to chronological rather than phenotypic age, did not. The inverse DNAmTL coefficient reflects its scaling rather than contrary biology, since DNAmTL decreases with age, so both the shorter DNAmTL with early menopause and its inverse association with mortality are directionally consistent with accelerated aging. Estrogen withdrawal and ovarian aging have been linked to metabolic, vascular, and immune remodeling, captured more directly by second-generation and pace-of-aging measures.^8,11,22^

The genetic analyses add nuance. ANM and EAA PRSs were inversely correlated, and genetically predicted later ANM was associated with lower epigenetic aging, mirroring the phenotype-level association; IV models were directionally consistent in both directions. The ANM-GrimAgeAccel association was notably robust, strengthening under pleiotropy-robust models, whereas HannumAgeAccel and PhenoAgeAccel attenuated with directions preserved. SNP-level Mendelian randomization and colocalization did not support widespread sharing of single causal variants,^21^ with most regions favoring distinct variants apart from chr6:29.98–30.98 Mb for ANM and HannumAgeAccel. The evidence is therefore most consistent with shared polygenic architecture and correlated pleiotropy rather than a simple causal chain, although limited instrument strength may contribute to the null SNP-level results: the published epigenetic age acceleration GWAS are considerably smaller than those for age at natural menopause, and their limited statistical power is a likely reason the colocalization analyses were not more compelling. Menopause timing and DNAm-based aging may thus partly reflect common biology related to ovarian aging, endocrine regulation, genome maintenance, and systemic aging.^9,10,12^

At the molecular marker level, MHT did not recapitulate the aging-marker pattern seen for menopause timing: broad MHT categories were unassociated with most EAA measures, yet current use showed much stronger EWAS and immune-marker signals than AAM. Genome-wide effect sizes for AAM and current MHT were essentially uncorrelated and only 57% of suggestive CpGs were directionally concordant, indicating largely non-overlapping CpG sets, with the same pattern in circulating cytokines and chemokines. Exogenous hormone exposure therefore appears to act more on immune and inflammatory pathways than on the aging metrics tied to menopause timing. Changes seen during current use persisted, attenuated, among former users, indicating a dampened, memory-like signature detectable a mean of approximately 6 years after stopping.

PLCO and the Women’s Health Initiative (WHI) enrolled women of similar ages in the 1990s. In our study, ever MHT use under pre-21st-century standards of care was associated with up to 30% lower BC-specific mortality, despite 19% higher HR+ BC incidence. The incidence excess was confined to women with natural menopause and was larger for estrogen plus progestin than for estrogen-only therapy, mirroring the divergence between the two WHI arms, whereas the mortality and survival advantages were present for both regimens and for all durations of use. This contrasts with current USPSTF recommendations, heavily influenced by the increased incidence observed among women with an intact uterus receiving combined estrogen-progestin therapy in the WHI,^7,28^ where many participants initiated therapy well beyond the transition. The FDA drew the same distinction between incidence and mortality in removing the boxed warnings, citing the absence of any trial showing increased breast cancer mortality with MHT and estimating that decades of blanket warnings had withheld benefit from some 50 million women.^7^ Our findings also contribute to the debate over whether earlier menopause drives biological aging, marks pre-existing susceptibility, or both.^12,21^ The genetic findings suggest shared architecture and correlated pleiotropy, supporting earlier menopause at least the genetically regulated portion as a marker of susceptibility, whereas our previous studies showing markedly stronger lung morbidity and mortality associations for bilateral oophorectomy (primarily due to benign gynecologic conditions such as ovarian cysts, benign tumors, endometriosis, and uterine fibroids, or risk-reducing bilateral salpingo-oophorectomy) than natural menopause support a causal contribution of estrogen loss.^1^ Clarifying their relative importance will require longitudinal studies with repeated molecular assessment.

Several limitations warrant consideration. First, it is a secondary analysis of an existing cohort: the serum immune and cytokine panel was assayed under the original EEMS substudy design, so the choice of analytes was fixed by that prior work. Harmonization across source substudies, fixed-effect meta-analysis, and consistent quality-control and lower-limit-of-detection filtering improve the credibility of the pooled estimates, but the panel itself was not under our control. Second, PLCO is an observational cohort, and residual or unmeasured confounding cannot be fully excluded. We applied causal-inference epidemiologic methods rigorously and, where germline instruments were available, used Mendelian randomization and statistical colocalization to provide instrumental-variable support; these approaches strengthen causal interpretation but do not replace the evidence a randomized trial would provide. Third, blood was drawn at a single timepoint, so we cannot establish the temporal ordering of hormone therapy and epigenetic aging. Distinguishing whether hormone therapy alters the epigenetic aging trajectory or instead marks an underlying one would require paired pre- and post-initiation samples.

In conclusion, counselling based on incidence alone is insufficient, since a shift toward hormone-receptor-positive disease can coexist with lower breast cancer mortality; because these exposures predate contemporary formulations, this argues for re-examining the balance in modern regimens rather than extrapolating to them. Earlier menopause marks systemic aging detectable more than a decade later and carries prognostic information after breast cancer. Resolving its causal role will require validation in newer and younger cohorts using contemporary hormone therapy, longitudinal cohorts with repeated molecular measurement and biospecimen collection around the menopausal transition, better-powered genome-wide studies of epigenetic aging, and replication in more diverse populations.

## Data Availability

PLCO data is under controlled access. Data access was approved under request PLCO-981 by the National Cancer Institute Cancer Data Access System on 24 June 2022 and under project ID 35044 by the database of Genotypes and Phenotypes (dbGaP) on 17 June 2024. Access to raw DNA methylation data is through dbGaP accession: phg002226.v1.

## Author contributions

SL conceptualized this project, requested the PLCO data from The Cancer Data Access System (PLCO-981) and supervised the data analyses. TZ conducted data analyses and summarized the results in tables and figures. TZ and SL interpreted the results and drafted the manuscript. VSP, KEZ, YL, KKL,YG, CIA, and LSC critically edited the manuscript. SL is responsible for the overall content as a guarantor.

## Data availability

PLCO data is under controlled access. Data access was approved under request PLCO-981 by the National Cancer Institute Cancer Data Access System on 24 June 2022 and under project ID 35044 by the database of Genotypes and Phenotypes (dbGaP) on 17 June 2024. Access to raw DNA methylation data is through dbGaP accession: phg002226.v1. Analysis codes will be made publicly accessible upon publication of this work at: https://github.com/zhaiting/PLCO.

