## Supplementary Materials for "Menopause timing, hormone therapy, epigenetic aging, and breast cancer outcomes: a secondary analysis of the PLCO Cancer Screening Trial"

### Supplementary Methods

#### Study Population

The study population was drawn from the Prostate, Lung, Colorectal and Ovarian (PLCO) Cancer Screening Trial. Data access was approved under request PLCO-981 by the National Cancer Institute Cancer Data Access System on 24 June 2022 and under project ID 35044 by the database of Genotypes and Phenotypes on 17 June 2024.

The original breast cancer cohort included 78,209 eligible female participants with trial data of cancer incidence from 1993 to 2009 and extended follow-up cancer registry data through 2020. A nested case-control study of breast cancer comprising 1,568 women had peripheral blood DNA methylation measured from baseline blood buffy coat samples collected prior to cancer diagnosis for cases.[1] It included 721 incident invasive primary breast cancer cases (ICD-9 174.0-174.9). The 847 controls were frequency matched on age at random assignment (5-year intervals), and fiscal year of randomization (pre/post 10/1/1997). All controls were alive and had no history of cancer as of the date of diagnosis for the matched case. Of these, 51 participants had poor methylation quality and were excluded, yielding 1,517 participants eligible for analysis. Participants missing the primary exposure: AAM (n=14) were removed (a further 3 women with ambiguous reporting of MHT use were retained but treated as missing in MHT-adjusted analyses). The final analytical cohort consisted of 1,503 women, all of self-reported White, non-Hispanic ancestry—the only race/ethnicity represented in the EPIC subset (**Figure 1**).

AAM was collected via baseline self-administered questionnaires using prespecified categorical ranges and harmonized into a five-level ordinal variable: < 40, 40–44, 45–49, 50–54, and ≥ 55 years. Menopause type (natural, surgical, and drug therapy) and MHT use (current, former, and never use) were obtained from baseline and confirmed via follow-up questionnaires. Participant characteristics were extracted from baseline questionnaires.[2]

#### DNA Methylation Processing

Raw DNA methylation data were generated using the Illumina Infinium MethylationEPIC BeadChip (EPIC v1). Raw EPIC array signals were preprocessed by the *minfi* R package, following standard quality control and normalization procedures. Briefly, samples with low median signal intensity (<10.5), high detection failure rates (more than 1% of probes with detection $P \geq 0.01$), or excessive intermediate β-values (more than 30% between 0.45–0.55) were excluded. Background correction and dye-bias adjustment were performed using the ssNoob method, followed by quantile normalization to reduce technical variation. Probes were further filtered to remove those with poor detection performance, known SNP overlap, or cross-reactivity. After quality control and filtering, a total of 824,126 probes were retained for downstream analyses. Blood cell-type proportions (CD4+ T cells, CD8+ T cells, B cells, NK cells, monocytes, and granulocytes) were estimated using reference-based deconvolution using the estimateCellCounts2 function within the *FlowSorted.Blood.EPIC* R package.

#### Epigenetic Age Estimation

DNA methylation–based epigenetic age was computed using the online platform hosted by the Clock Foundation (https://dnamage.clockfoundation.org). Six DNAm aging metrics were evaluated in total: four established epigenetic age clocks, Horvath, Hannum, PhenoAge, and GrimAge, together with DNAm-based telomere length (DNAmTL) and DunedinPACE, each described in turn below. EAA was defined as the residual from regressing DNAm age clocks on chronological age, adjusting for estimated blood cell-type composition (**Figure S1A**).[3,4] DNAm-based telomere length (DNAmTL) was estimated using the epigenetic telomere length predictor developed by Lu et al., which captures the replicative aging dimension of cellular senescence from blood DNAm profiles.[5] DunedinPACE, a pace-of-aging estimator trained on longitudinal within-person change, was computed from the same DNAm data using the DunedinPACE R package; unlike cumulative clocks that estimate total epigenetic wear, DunedinPACE captures the current rate of biological aging at the time of blood draw.[6]

**Epigenome-Wide Association Study (EWAS)**

EWAS was performed using limma with empirical Bayes shrinkage. M-values (logit-transformed β-values) were used as the outcome. The main predictor was AAM (modeled as a numeric ordinal variable from 1 to 5). Models were adjusted for chronological age, BMI, education, MHT use, menopause type, cancer diagnosis, cell type compositions, and array-related technical covariates (slide and position). Since 824,126 tests were performed across CpG probes, Benjamini-Hochberg (BH) false discovery rate (FDR) correction was applied. Because FDR correction at the probe level can be conservative in EWAS, Manhattan plots and locus prioritization additionally used a predefined raw p-value threshold of $5 \times 10^{-6}$ to highlight suggestive associations. All analyses showed minimal genomic inflation ($\lambda < 1.05$). Parallel EWAS models were fitted with MHT use (current vs never; former vs never) as the predictor, using the same M-value outcomes and BH-FDR multiple-testing correction, with AAM replacing MHT use in the covariate set.

Gene set enrichment analysis was conducted using *methylGSA*, which accounts for variable CpG–gene mapping and probe-count bias as previously described.[7,8] We applied the p-value based mRRA GSEA framework using raw CpG-level EWAS p values as input, given the limited number of CpG sites reaching FDR significance at the probe level. Pathway-level significance was assessed using FDR correction within each enrichment analysis. Analyses focused on MSigDB Hallmark pathways (v7.1) and KEGG pathways.

**Polygenic Risk Score**

Genome-wide genotype data for PLCO participants were obtained from dbGaP (accession phg001677). Genotyping was conducted on multiple array platforms across ancestry groups and imputed using the TOPMed reference panel (release 5b). We restricted analyses to participants of European (EUR) ancestry, the largest available subset and the closest match to the genetic ancestry of the analytic cohort, for phenotype-level association testing. Detailed genotyping and preprocessing procedures done at the PLCO have been described previously.[9] We applied additional variant-level filtering using PLINK2 (v2.0). Variants were excluded if they had minor allele frequency (MAF) < 0.01, genotype call rate < 98%, or imputation quality (Mach-$R^2$) < 0.8. After filtering, 8,098,937 variants across chromosomes 1–23 were retained for downstream analyses.

Polygenic risk scores (PRS) for age at natural menopause (ANM) were derived using summary statistics from the largest European-ancestry GWAS to date by Ruth et al.[10] using autosomal variants. PRSs were constructed using LDpred2 (auto model), with linkage disequilibrium (LD) estimated directly from the European-ancestry PLCO subset (N = 100,445). Analyses were restricted to HapMap3 variants to ensure stable LD estimation. Of the quality-controlled autosomal variants in PLCO (7,878,706 SNPs), 720,612 HapMap3 variants remained after matching to GWAS summary statistics and were used for PRS derivation. Resulting PRSs were standardized to mean 0 and standard deviation 1. PRS of ANM explained 9.9% variations in ANM phenotype within the European-ancestry PLCO subset, close to reporting in the original GWAS (10.1%) (**Figure S2A**).

PRSs for EAA were derived using GWAS summary statistics from McCartney et al.[11] PRS construction followed the standard LDpred2 auto workflow restricted to autosomal HapMap3 variants, with LD estimated from the European-ancestry PLCO subset. Scores were standardized to mean 0 and standard deviation 1. PRS of epigenetic ageing metrics explained variations for 1–3% within the European-ancestry PLCO subset, consistent with reporting in the original GWAS.

Principal components (PCs) were derived from genome-wide genotype data using PLINK. Based on inspection of the eigenvalue distribution and scree plot, the first eight PCs were retained for adjustment in the genetic analyses (**Figure S2D**). These components together explained approximately 67% of the total genetic variance.

**Instrumental Variables Analyses Via Structural Equation Modeling**

To evaluate directional associations between ANM and EAA, we implemented PRS-based instrumental-variable (IV) models using structural equation modeling (SEM) using the lavaan R package among a subset of women who experienced natural menopause. Effect estimates reflect one standard deviation (SD) increases in genetic liability.

In the forward direction (ANM to epigenetic aging metrics), ANM was modeled as the ordered 5-level endogenous variable. Instrument strength was assessed by regressing this ordinal variable on the ANM PRS and genetic principal components (PC1–PC8), yielding the first-stage coefficient (**a**). Each epigenetic aging metrics (HorvathAgeAccel, HannumAgeAccel, PhenoAgeAccel, GrimAgeAccel, DunedinPACE, and DNAmTL) was then regressed on genetically predicted later ANM and PC1–PC8 to estimate the second-stage association (**b**). The product (**a**x**b**) was reported as the reduced-form effect, reflecting the association between the genetic instrument and the outcome mediated through ANM. Primary analyses enforced the IV exclusion restriction by constraining the direct path from the ANM PRS to the EAA outcome to zero. To assess robustness to potential horizontal pleiotropy, sensitivity analyses additionally allowed a direct PRS to outcome path (**c**), relaxing the exclusion restriction. All forward-direction models were fitted using the weighted least squares mean- and variance-adjusted (WLSMV) estimator with ANM specified as an ordered endogenous variable.

In the reverse direction (EAA to ANM), analogous SEM-based IV models were fitted using PRSs for EAA (HannumAgeAccel, PhenoAgeAccel, and GrimAgeAccel) as instruments. In these models, the EAA metric was treated as the exposure, and ANM was modeled as the ordered outcome. Instrument strength (a), directional association (b), and reduced-form effects were estimated in the same manner as in the forward analyses. Sensitivity analyses allowing direct PRS-to-ANM paths were similarly conducted to probe robustness to pleiotropy.

**SNP-level analyses using two-sample MR and colocalization**

To complement PRS-based instrumental variable analyses and further interrogate genetic relationships at the variant level, we conducted SNP-level analyses using two-sample Mendelian randomization (MR) and locus-specific colocalization based on GWAS summary statistics for ANM and EAA measures used in PRS construction.

Primary MR analyses were conducted using CAUSE (Causal Analysis Using Summary Effect estimates), a Bayesian framework that leverages genome-wide summary statistics and explicitly models correlated horizontal pleiotropy.[12] CAUSE jointly estimates causal and shared pleiotropic effects and accounts for potential sample overlap between exposure and outcome GWAS, providing robustness in settings where genetic correlation between traits is expected. Summary statistics were harmonized prior to analysis, and default prior specifications were used. Evidence for a causal effect was evaluated by comparing the fit of the causal model against the sharing (pleiotropic) model using expected log pointwise predictive density (ELPD).

As complementary analyses, we applied conventional two-sample MR estimators implemented in the *TwoSampleMR* R package,[13] including inverse-variance weighted (IVW), MR-Egger, weighted median, simple mode, and weighted mode methods, to assess consistency across modeling assumptions. For these analyses, genetic instruments were selected using a genome-wide significance threshold ($p < 5\times10^{-8}$), clumped to ensure independence (10 Mb window, $r^2 < 0.001$), and restricted to variants with F-statistics greater than 10 to minimize weak-instrument bias. To further evaluate potential horizontal pleiotropy, we conducted MR-PRESSO (Mendelian Randomization Pleiotropy RESidual Sum and Outlier) analyses to identify outlier variants and test for distortion in causal estimates due to pleiotropic effects.[14] MR-PRESSO was implemented with 20,000 permutations (NbDistribution = 20,000) and a significance threshold of 0.05.

To evaluate whether observed associations between ANM and EAA could be driven by shared causal variants at specific loci, we performed colocalization analyses using the *coloc* R package with the SuSiE (Sum of Single Effects) fine-mapping framework.[15] This approach relaxes the assumption of a single causal variant per locus and allows for multiple causal signals in regions with complex linkage disequilibrium (LD). Candidate loci for colocalization were prioritized based on joint evidence of association in both traits: lead SNPs were identified using the minimum p-value across ANM ($P < 1x10^{-5}$) and EAA ($P < 1x10^{-3}$) GWAS summary statistics, followed by LD-based pruning to define approximately independent signals. Genomic regions were defined as ±500 kb windows centered on lead SNPs, and only regions with sufficient overlapping SNP coverage (≥200 SNPs) were retained for analysis. LD matrices were estimated using an independent European-ancestry reference panel from the 1000 Genomes Project (Phase 3, EUR) with PLINK, restricting variants to minor allele frequency ≥1% to ensure stable LD estimation. Summary statistics for ANM and EAA were harmonized to the LD reference panel, and allele alignment was verified prior to fine-mapping. Within each region, SuSiE fine-mapping was performed separately for ANM and EAA using default credible set coverage (0.95). The maximum number of effects was set to L = 10 for ANM and L = 3 for EAA, reflecting differences in GWAS sample size and expected locus complexity. Colocalization posterior probabilities were computed for all pairs of SuSiE signals across traits and summarized under the five standard hypotheses: no association (H0), association with ANM only (H1), association with EAA only (H2), distinct causal variants (H3), and a shared causal variant (H4). Evidence for colocalization was inferred only when posterior support for H4 substantially exceeded that for H3.

**Immune Marker Analysis**

Immune and inflammation markers were obtained from five PLCO Etiologic and Early Marker Studies (EEMS) case-control studies (lung, second lung, NHL, ovarian, and endometrial cancer; https://cdas.cancer.gov/learn/plco/mips/). EEMS was designed to identify etiologic and early-detection blood-based immune and inflammation markers that precede cancer diagnosis, using pre-diagnostic serum in nested case-control studies within PLCO. The two pilot studies and the upper GI study were excluded due to sparsity of analytes and small sample size. Analytes were measured primarily via multiplex Luminex bead–based assays (Millipore), with single-plex immunoassays used for selected markers. Concentrations were derived via 4- or 5-parameter logistic calibration curves according to laboratory protocols. Within each study, matched pairs were assayed together, with blinded QC replicates included in each batch. LLOD is reported in previous literature and not included in the provided dataset, but the QC flag was included for identification of values outside LLOD.[16,17]

Data from the five studies were pooled (n=3,677 all-sex; after removing 62 duplicate measures collected post baseline). We restricted to female-only participants, yielding a pooled female EEMS marker-QC cohort in which per-marker denominators range up to N=1,992 depending on substudy availability. After harmonizing marker definitions and measurement units, markers with more than 80% below the LLOD (n=18) and more than 90% missing values (n=12) across this marker-QC cohort were excluded by upstream QC, yielding 74 analytes (**Table S1**); among these, 6 were available only in the endometrial substudy. For marker-level regression analyses, we further restricted to participants with complete exposure and covariate data, yielding an analytic sample of n=1,933 women. Among the 74 QC-retained markers, 48 had at least 50% non-missing values in this analytic sample and were used for regression testing; the remaining 26 markers were largely restricted to study-specific panels (e.g., adipokines and metabolic peptides assayed only in the endometrial substudy, or T-cell cytokines measured in fewer than 3 of 5 substudies) and were excluded to avoid unstable per-marker fits. All marker regressions included source-substudy fixed effects to account for study-specific assay calibration; we additionally performed fixed-effect meta-analyses across substudies for each retained marker to evaluate between-study heterogeneity (**Table S2**). Marker concentrations were log-transformed and standardized before analysis. Associations between AAM and each marker were estimated using linear models adjusting for age, BMI, MHT use, and case-control status, with additional models including menopause type. BH FDR was applied to adjust for multiple testing. Parallel models were fitted with current MHT use (vs never) as the predictor, substituting AAM into the covariate set.

**Statistical Analysis**

Descriptive statistics were summarized as means (standard deviations) for continuous variables and frequencies (percentages) for categorical variables. Between-group comparisons used two-sided t-tests and $\chi^2$ tests.

Associations of AAM and MHT use with BC incidence, mortality, and overall survival among BC cases were evaluated in the full PLCO female screening cohort (N=78,209) using Cox proportional hazards models. BC incidence was defined as time from enrollment to first incident BC (pooled in situ and invasive), censored at the end of PLCO BC ascertainment (through 2020); BC-specific mortality was defined as time from enrollment to BC death adjudicated by the PLCO Death Review Committee, censored at the mortality follow-up cutoff (through 2022); overall survival among BC cases was time from BC diagnosis to death from any cause or last contact, censored at the mortality follow-up cutoff (through 2022). MHT use includes ever versus never, number of years taking female hormones (0 [never], ≤1 Year, 2-3 Years, 4-5 Years, 6-9 Years, and 10+ Years), and MHT regimens (estrogen pill, progestin pill, estrogen/progestin pill, cream of estrogen, progestin, or estrogen/progestin, unknown MHT). MHT regimens were defined using Baseline Questionnaire and Supplemental Questionnaire. We fitted an age-adjusted model (Model 1) and a multivariable model (Model 2) additionally adjusting for BMI, education, MHT use or menopause timing as appropriate, menopause type, and smoking status; for overall survival among BC cases, both Model 1 and Model 2 additionally adjusted for BC stage at diagnosis. Participants with a missing value on a categorical covariate were retained by assigning them to an explicit unknown category for that covariate rather than excluded, preserving the full multivariable sample; in the subtype-stratified analyses, competing- and unknown-subtype cases were censored at their event time. Proportional hazards assumptions were assessed using Schoenfeld residuals and time-interaction terms. In the EPIC methylation subset, Cox proportional hazards models were also used to evaluate each epigenetic aging metric as an independent predictor of overall survival among BC cases, with hazard ratios expressed per one standard deviation increase.

AAM was treated both as an ordinal 5-level exposure for statistical power and binary by comparing early menopause (< 45 years) to the reference category (≥45 years) for interpretability in survival analyses.[2] For EAA measures, associations of AAM and MHT use with epigenetic aging metrics were evaluated using linear models. These models were fitted as either crude models without covariate adjustment or adjusted for body mass index (BMI), education, MHT use (for AAM models) or AAM (for MHT models), menopause type, and BC status; chronological age and cell-type compositions were additionally included for models of DNAmTL and DunedinPACE, as these covariates were not accounted for during their derivation. Analogous models were used to examine associations of AAM and MHT use with DNAm-estimated blood cell-type proportions. Effect modification was evaluated through stratified analyses by menopause type (natural and surgical), smoking status (never and ever smoker), and BC status (case and control), which were predefined as important factors that might have heterogeneous relationships, based on prior literature and study design.[2,18] Ever MHT use, MHT duration, and MHT regimen × clinical endpoint analyses were additionally stratified by menopause type (natural versus surgical) in exploratory analyses; women with radiation-induced or drug-induced menopause and those with unknown menopause type were not included in the stratified analyses. MHT regimen was taken from the supplemental questionnaire item on the type of therapy first used, aligned with the baseline questionnaire, and was available for the subset of participants who completed that questionnaire; ever users whose regimen was not reported or who answered "not sure" were retained as an explicit unknown-regimen group.

We evaluated potential mediation of the AAM–survival association through epigenetic aging metrics using two complementary approaches. First, sequential covariate adjustment assessed the degree of attenuation when aging biomarkers were added individually to the multivariable Cox model.[18] The Model 2 covariate set served as the base for all mediation analyses, with each aging biomarker added as an additional covariate (not substituted for the multivariable adjustment); attenuation of the exposure HR therefore reflects the incremental contribution of the biomarker beyond demographic and reproductive covariates. Second, formal regression-based causal mediation estimated total natural indirect effects (TNIE) through each biomarker individually, using the CMAverse R package with Cox proportional hazards models for time-to-event outcomes.[19] Direct, indirect, and total effects, as well as the proportion mediated, were estimated with 2,000 bootstrap resamples following a previously described framework. Mediation analyses were restricted to BC cases in the EPIC methylation subset.

To assess the degree of concordance between menopause timing and MHT use at the molecular level, genome-wide Pearson correlation coefficients were computed between EWAS effect estimates for AAM and

MHT use, and between immune marker effect estimates for AAM and MHT use. Directional consistency was quantified as the proportion of suggestive CpGs or markers showing same-direction effects across both exposures. Pearson correlation of EWAS effect estimates for former MHT use against current MHT use was also computed at CpGs significant for current MHT and across all CpGs genome-wide. Correlation analyses between PRS for ANM and EAA metrics were conducted using Pearson correlation coefficients.

All statistical tests were two-sided and $p<0.05$ was considered significant unless otherwise noted. Where applicable, multiple testing was controlled using FDR correction within analytic domains (EWAS, pathway enrichment). All analyses were performed in R (v4.4.1).

**Supplementary Tables**

Table S1. Immune marker panel quality control results.

Table S2. Serum immune marker associations with age at menopause and hormone therapy use, with fixed-effect meta-analysis across source substudies.

Table S3. Ordinal age at menopause in relation to breast cancer and mortality endpoints in the full PLCO cohort.

Table S4. Ever hormone therapy use, duration of hormone therapy use, and hormone therapy regimen in relation to breast cancer and mortality endpoints, by menopause type.

Table S5. Molecular subtype-stratified analyses for ever hormone therapy use and early menopause.

Table S6. Temporal gap between age at menopause and DNAm blood draw, by age-at-menopause category.

Table S7. Epigenetic aging metrics by exposure category and subgroup.

Table S8. DNAm-estimated immune cell proportions by age at menopause and hormone therapy status.

Table S9. Epigenetic aging metrics in between breast cancer cases and controls.

Table S10. Sensitivity analyses of PRS-based instrumental variable models.

Table S11. SuSiE-based colocalization between age at natural menopause and epigenetic aging GWAS loci.

Table S12. Two-sample Mendelian randomization analyses of age at natural menopause and epigenetic aging acceleration.

Table S13. Top 20 CpGs associated with age at menopause in the EPIC methylation subset.

Table S14. FDR-significant CpGs from EWAS of current hormone therapy use.

### Supplementary Figures

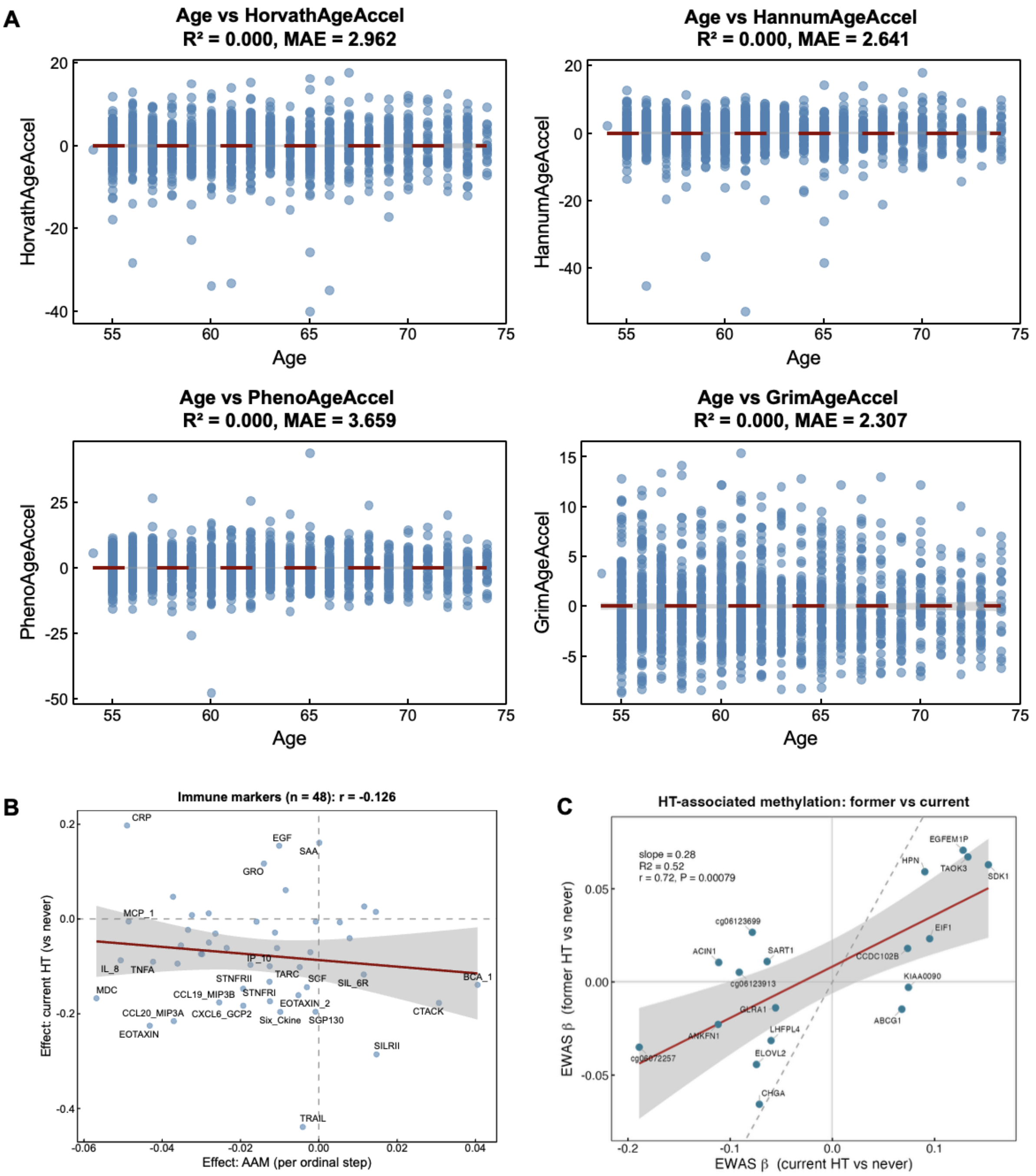


**Figure S1.** (**A**) Scatter plots of chronological age versus epigenetic age acceleration (EAA) measures, including HorvathAgeAccel, HannumAgeAccel, PhenoAgeAccel, and GrimAgeAccel. Epigenetic age acceleration was defined as the residual from regressing DNAm age on chronological age, adjusting for estimated blood cell composition. Red horizontal lines indicate zero acceleration. $R^2$ and mean absolute error (MAE) are shown for each model. (**B**) Immune marker associations: age at menopause vs hormone therapy. Scatter plot of paired effect estimates for 48 serum immune markers comparing AAM (x-axis) and current HT (y-axis) associations. (**C**) EWAS effect estimates (β) in former vs current HT users at the 18 current-HT FDR<0.05 CpGs. X-axis, current HT vs never; y-axis, former HT vs never. Red line, linear regression with 95% confidence band (grey); dashed grey line, y=x identity. Points labelled with mapped gene symbol or CpG ID.

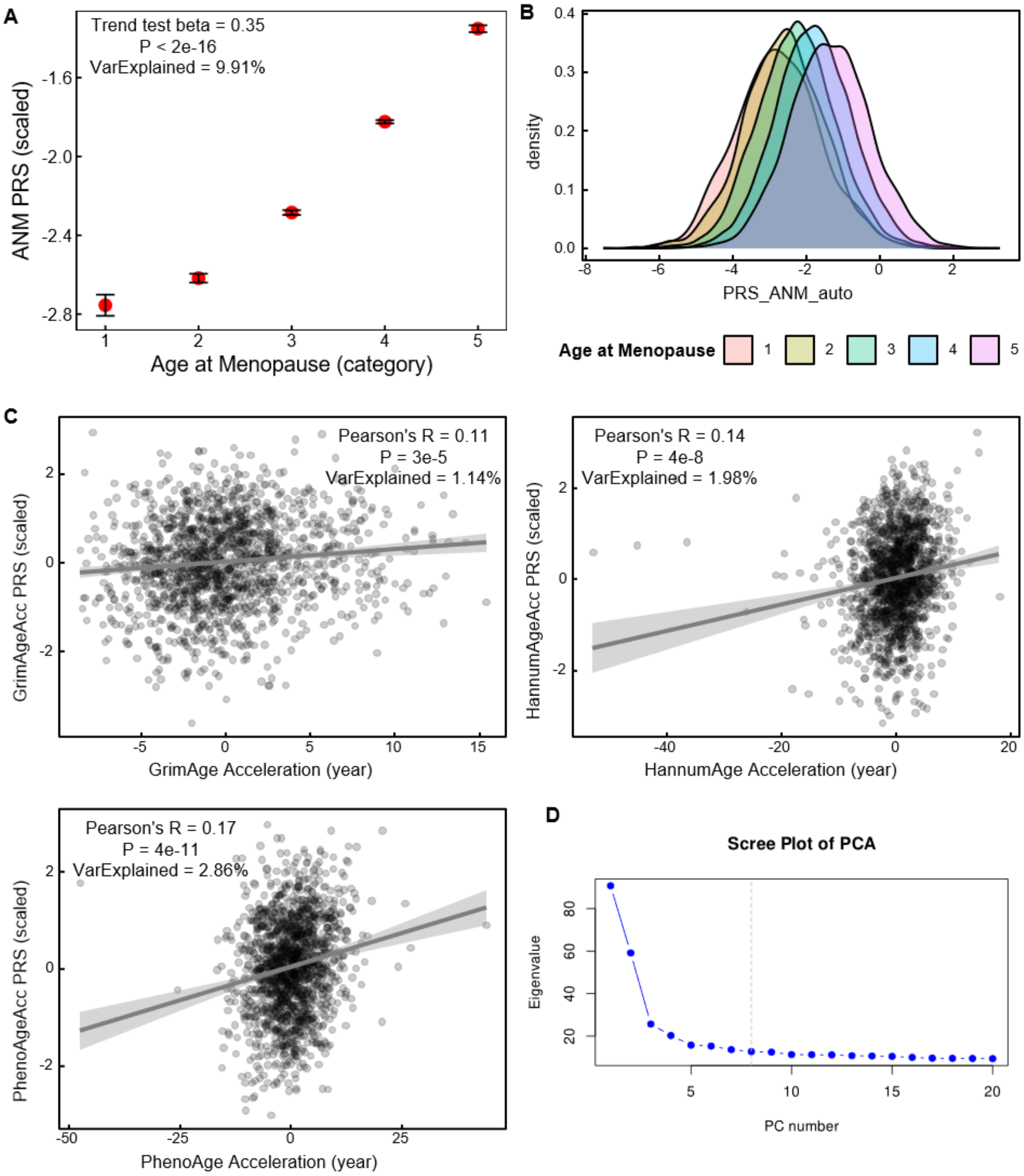


**Figure S2.** Polygenic risk scores for age at natural menopause and epigenetic age acceleration in the PLCO cohort. **(A)** Association between age at natural menopause (ANM; modeled as a five-level ordinal phenotype) and the standardized autosomal polygenic risk score for ANM (PRS_ANM_auto). Points represent mean PRS values for each ANM category, with error bars indicating standard errors. The linear trend test, corresponding P value, and proportion of variance explained are shown. **(B)** Density plots of PRS_ANM_auto stratified by ANM category, illustrating a monotonic shift toward higher genetic predisposition for later menopause across increasing phenotype categories. **(C)** Scatter plots showing associations between epigenetic age acceleration phenotypes and their corresponding polygenic risk scores: GrimAge acceleration vs GrimAgeAccel PRS, HannumAge acceleration vs HannumAgeAccel PRS, and PhenoAge acceleration vs PhenoAgeAccel PRS. Each point represents an individual participant. Solid lines indicate linear regression fits with shaded 95% confidence intervals. Pearson correlation coefficients (R), P values, and variance explained are reported. **(D)** Scree plot from principal component analysis (PCA) of genome-wide genotype data performed using PLINK. Eigenvalues are shown for the top principal components; the dashed vertical line indicates the number of PCs (8) retained for adjustment in genetic analyses.

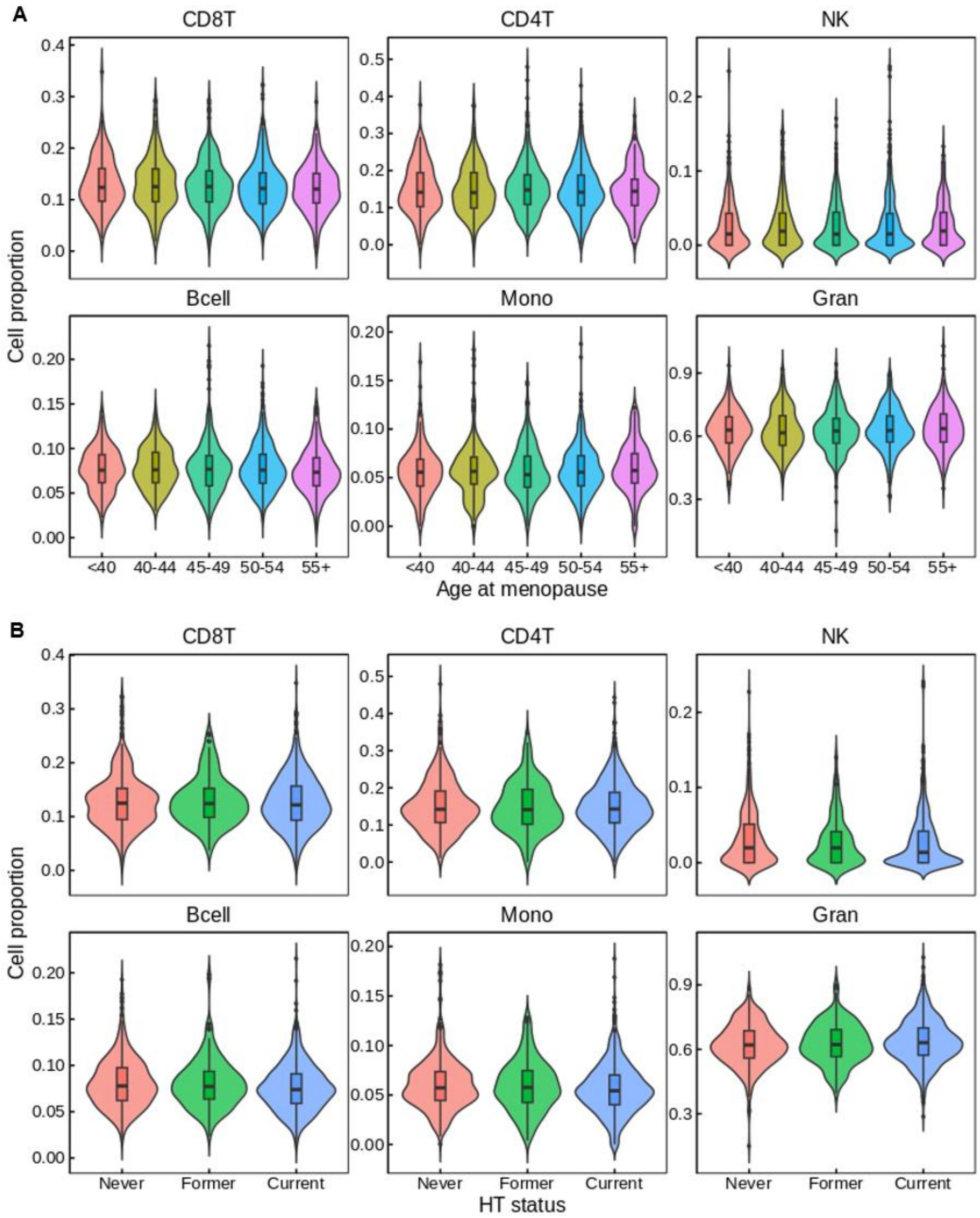


**Figure S3.** Blood cell composition by age at menopause and independence of epigenetic age acceleration from chronological age. (**A**) Violin plots showing estimated proportions of major blood cell types (CD8+ T cells, CD4+ T cells, NK cells, B cells, monocytes, and granulocytes) across categories of age at menopause (<40, 40−44, 45−49, 50−54, ≥55 years). Cell-type proportions were estimated from DNA methylation data using reference-based deconvolution. Boxes within violins indicate the median and interquartile range; points represent individual participants. (**B**) DNAm-estimated immune cell proportions by hormone therapy status. Violin plots showing Houseman deconvolution estimates for six major cell types by HT status (never, former, current).
